# Leveraging sequences missing from the human genome to diagnose cancer

**DOI:** 10.1101/2021.08.15.21261805

**Authors:** Ilias Georgakopoulos-Soares, Ofer Yizhar Barnea, Ioannis Mouratidis, Candace S.Y. Chan, Rachael Bradley, Mayank Mahajan, Jasmine Sims, Dianne Laboy Cintron, Ryder Easterlin, Julia S. Kim, Emmalyn Chen, Geovanni Pineda, Guillermo E. Parada, John S. Witte, Christopher A. Maher, Felix Feng, Ioannis Vathiotis, Nikolaos Syrigos, Emmanouil Panagiotou, Andriani Charpidou, Konstantinos Syrigos, Jocelyn Chapman, Mark Kvale, Martin Hemberg, Nadav Ahituv

**Affiliations:** Department of Bioengineering and Therapeutic Sciences, University of California San Francisco, San Francisco, California, USA; Institute for Human Genetics, University of California San Francisco, San Francisco, California, USA; Institute for Personalized Medicine, Department of Biochemistry and Molecular Biology, The Pennsylvania State University College of Medicine, Hershey, PA, USA; Department of Computer Science, Katholieke Universiteit Leuven, Leuven, Belgium; Evergrande Center for Immunologic Diseases, Harvard Medical School and Brigham and Women’s Hospital, Boston, USA; Department of Epidemiology and Biostatistics, University of California San Francisco, San Francisco, California, USA; Division of Gynecologic Oncology, University of California San Francisco, San Francisco, California, USA; Donnelly Centre for Cellular and Biomolecular Research, University of Toronto, Toronto, Canada; Department of Epidemiology and Population Health, Stanford University, Stanford, California, USA; Division of Oncology, Department of Medicine, Siteman Cancer Center, Washington University School of Medicine, St. Louis, Missouri; Siteman Cancer Center, Washington University School of Medicine, St. Louis, Missouri; Department of Biomedical Engineering, Washington University School of Medicine, St. Louis, Missouri; Division of Hematology/Oncology, Department of Medicine, University of California San Francisco, San Francisco, California, USA; Helen Diller Comprehensive Cancer Center, University of California San Francisco, San Francisco, California, USA; Department of Radiation Oncology, University of California San Francisco, San Francisco, California, USA; Department of Urology, University of California San Francisco, San Francisco, California, USA; Third Department of Internal Medicine, Sotiria Hospital, National and Kapodistrian University of Athens, School of Medicine, Athens, Greece; Department of Pathology, Yale School of Medicine, New Haven, CT, USA; Breast Oncology, Dana-Farber Brigham Cancer Center, Boston, MA, USA; Wellcome Sanger Institute, Hinxton, UK

**Author notes:** These authors contributed equally to the work (Ilias Georgakopoulos-Soares, Ofer Yizhar Barnea).

## Abstract

Cancer diagnosis using cell-free DNA (cfDNA) has potential to improve treatment and survival but has several technical limitations. Here, we show that tumor-associated mutations create neomers, DNA sequences 13-17 nucleotides in length that are predominantly absent from genomes of healthy individuals, that can accurately detect cancer, including early stages, and distinguish subtypes and features. Using a neomer-based classifier, we show that we can distinguish twenty-one different tumor-types with higher accuracy than state-of-the-art methods. Refinement of this classifier using a handcrafted set of kmers identified additional cancer features with greater precision. Generation and analysis of 451 cfDNA whole-genome sequences demonstrates that neomers can precisely detect lung and ovarian cancer with an area under the curve (AUC) of 0.93 and 0.89, respectively. In particular, for early stages, we show that neomers can detect lung cancer with an AUC of 0.94 and ovarian cancer, which lacks an early detection test, with an AUC of 0.93. Finally, testing over 9,000 sequences with either promoter or massively parallel reporter assays, we show that neomers can identify cancer-associated mutations that alter regulatory activity. Combined, our results identify a novel, sensitive, specific and simple diagnostic tool that can also identify novel cancer-associated mutations in gene regulatory elements.

## Introduction

Cancer is the second leading cause of death worldwide^1,2^. For most cancer types, survival is significantly higher if the tumor is detected at an early stage^3,4^. Currently, mass population screening is applicable only for breast and cervical cancers and utilizes physical tests like mammography and cytology screens. Detection for other cancer types, done both *en masse* and in a low and affordable resource setting, still poses a major challenge for the scientific and clinical communities^5^. In particular, a major hurdle is to identify reliable biomarkers for the detection of cancer at a presymptomatic stage. Detection at such an early stage would allow not only for improved survival but would also decrease treatment toxicity and provide an opportunity for providing personalized treatments.

Circulating cell-free DNA (cfDNA) is an emerging and promising resource for cancer diagnostics and prognostics^6–8^. It has a short life span (16 minutes to 2.5 hours), which makes it a highly temporal indicator of various processes occurring in the subject’s body. Due to advances in sequencing technologies, cfDNA can be rapidly analyzed at a relatively low cost. Analysis of circulating tumor DNA (ctDNA) has become a prospective minimally invasive tool to screen the population and to monitor patients already diagnosed with cancer. To distinguish cancerous cells, their tissue of origin, cancer type, minimal residual disease and other cancer features current technologies rely on sequencing to detect somatic mutations^9^ and epigenetic marks, such as DNA methylation or histone modifications that can determine the cancerous tissue of origin^10,11^. However, ctDNA still has many hurdles and caveats that need to be overcome^12^. Some of the major hurdles include: 1) cfDNA is fragmented (180-360 base pairs) making its collection and extraction challenging; 2) tumor-derived DNA makes up only a small portion of the total cfDNA (estimated to be around 0.4%), necessitating the need for extremely sensitive biomarkers that can detect the presence of cancerous cells; 3) prior knowledge of specific mutations or methylation marks is required for targeted screening, and consequently the main focus has been on coding mutations which only constitute a small fraction of mutations. 4) cfDNA mutations and epigenetic diagnosis is confounded by somatic alterations in white blood cells^13^; 5) the diagnostic techniques used to detect methylation or histone marks are technologically complex and have low sensitivity and specificity, in particular in early stages^6,14–16^. To provide the most optimal cancer treatment, it needs to be diagnosed at preliminary stages when the tumor is small (∼5mm in diameter). At these stages, the tumor produces minute levels of ctDNA that are difficult to detect using current methods^6^.

Nullomers are short DNA sequences (13-17 base pairs) that are absent from the human genome^17–19^. While the absence of nullomers could be due to chance, we and others have shown that a significant proportion of them is under negative selection pressure^18,19^, suggesting that they may have a deleterious effect on the genome. We have also shown that these sequences could be used as DNA ‘fingerprints’ to identify specific human populations^19^. As nullomers generally do not exist in a human genome, their appearance due to mutagenesis followed by clonal expansion could be exploited as a diagnostic method for diseases associated with a mutational burden, such as cancer.

Here, we set out to test whether nullomers could be used as a diagnostic tool to detect cancer and various additional tumor features. Throughout this manuscript, we refer to nullomers found in multiple tumor genomes as *neomers* to distinguish them from the more general category. We first analyzed The Cancer Genome Atlas (TCGA;^20^) database finding that neomers created by somatic mutations are able to detect cancer subtypes with higher accuracy than leading methods^21^ as well as additional cancer features. Further analyses of cfDNA whole-genome sequencing (WGS) datasets found that these neomers can also be used to detect cancer subtypes. Using WGS on cfDNA from 451 individuals with ovarian or lung cancer and normal controls found an enrichment for neomers. Our neomer-based cancer detection models had AUCs of 0.93 for lung and 0.88 for ovarian cancers, and an AUC of 0.94 for lung and 0.93 for ovarian cancer at early stages. This is particularly noteworthy for ovarian cancer as currently there is no effective screening test^22^, with most women being diagnosed at later stages, IIIC or IV, where 5-year survival rates are 39% and 17%, respectively^23^. Finally, utilizing both promoter assays and a massively parallel reporter assay (MPRA), we show that neomers alter regulatory activity and can be used to detect cancer-associated mutations in gene regulatory elements. Combined, our results show that neomers can be used as a rapid, sensitive, specific and simple cancer diagnostic tool and also aid in the identification of gene regulatory mutations associated with cancer.

## Results

### Annotation of mutations that lead to nullomers

As cancer is associated with a large number of somatic DNA mutations, we investigated if they result in the resurfacing of nullomers (**Fig. 1a**). Using our previously characterized human nullomers^19^, we analyzed WGS results from 2,577 patients across 21 cancer types from TCGA^24^ for resurfacing nullomers (**Extended Data Fig.1a**). We focused on 16bp nullomers, as it is the shortest length where we detect a sufficient number of nullomers per patient, with the human reference genome having only 37.24% of all possible 16mers. The majority of the 44,599,472 single nucleotide substitutions give rise to multiple nullomers, allowing us to identify 213,164,038 resurfacing nullomers across all cancer types. Furthermore, we identified 2,470,091 nullomers resulting from short insertions and deletions (1-100 base pairs). The median number of nullomers created by each substitution was two and for indels four (**Extended Data Fig.1b-c**). On average, 58.29% of substitutions in a patient resulted in one or more nullomers, with 2.1% of the nullomers residing in coding regions. The median number of nullomers found across cancer patients was 9,107 (**Fig. 1b-c**) and the number of nullomers was directly proportional to the number of mutations (**Extended Data Fig.1d**). As mutations were identified by comparisons to healthy tissues, we did not filter for common variants that could result in nullomers^19^.

**Fig. 1:**
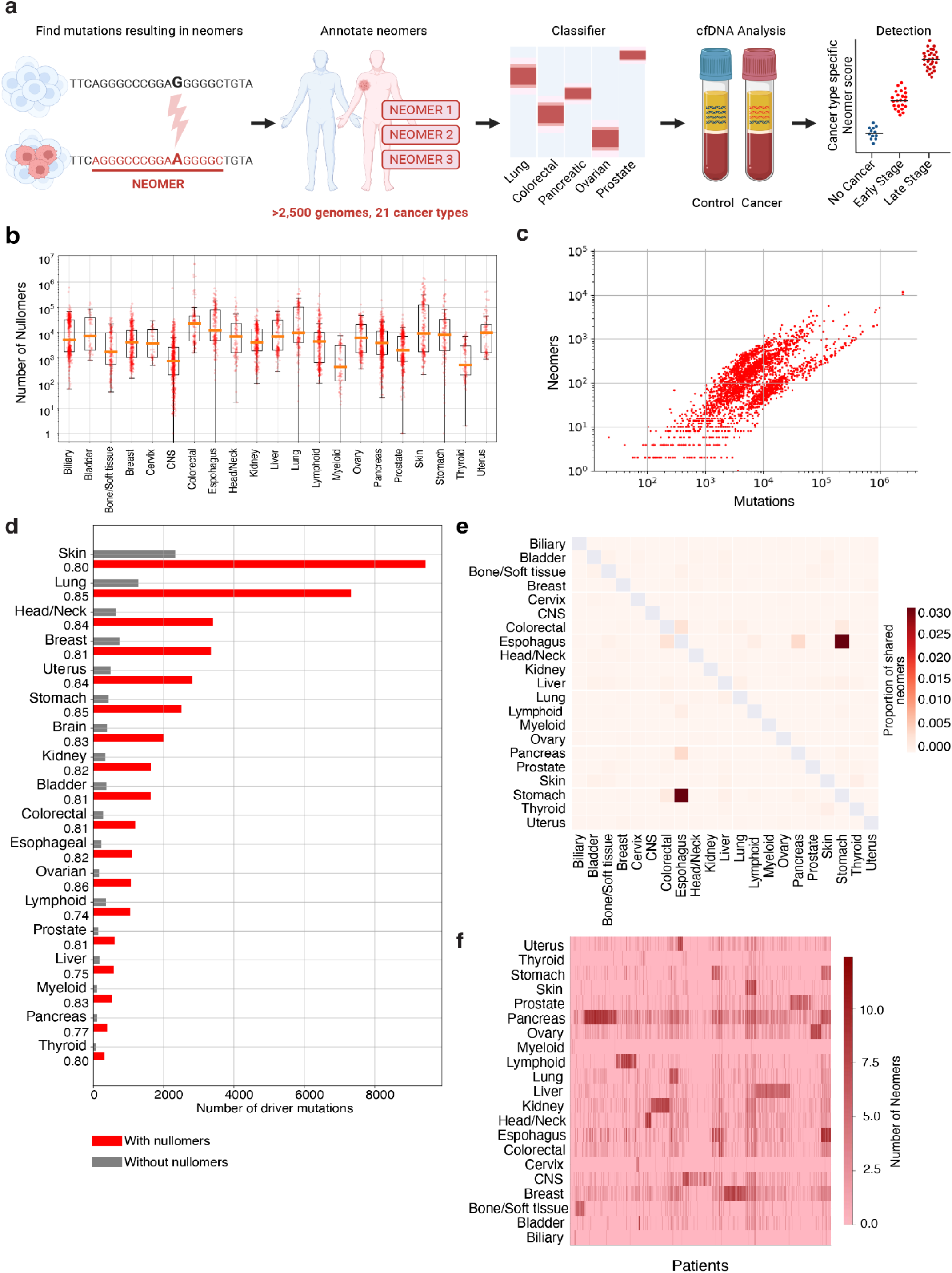
Neomers can detect cancer tissue of origin. **a**, Schematic overview of neomer cancer diagnostic pipeline. **b**, Number of neomers per patient sample across tissues. Each dot represents a patient sample. **c**, Number of neomers and the number of substitutions for 2,577 patients (Spearman’s rho = 0.75). **d**, Identification of nullomers at driver mutation sites across cancer tissues. The majority of cancer driver mutations result in the resurfacing of nullomers. The proportion of mutations that cause the resurfacing of nullomers is indicated under each cancer type.**e**, Heatmap showing the Jaccard index for the overlap of neomer sets associated with different cancer types. **f**, Heatmap showing the occurrence of neomers across patients for each cancer type. Each row represents a cancer type and each column a patient. The intensity of the heatmap (log2-scale) shows the number of neomers for each tissue set.

To further prioritize nullomers that could be used as cancer biomarkers, we focused on the subset of nullomers that are recurrent, i.e. those found in more than one patient for a specific cancer type, termed hereafter as *neomers*. The number of neomers was proportional to the total number of mutations (**Fig. 1c, Supplementary Table 1**). As both the number of patients per cancer type and the mutational load varied, the median number of neomers for each tissue type ranged from 0-98. Analysis of the most frequent neomers revealed several previously known cancer-associated driver mutations (**Table 1**). For example, some of the most recurrent coding neomers were the result of either the Gly12Asp, Gly12Val or Gly12Cys missense mutation in the KRAS proto-oncogene GTPase (KRAS), which are known to make up 80% of cancer-associated KRAS mutations and lead to KRAS being constitutively active^25,26^. Although KRAS mutations have been associated with several cancer types, 190/215 (88%) of these mutations were found in pancreatic cancers. Several frequently occurring coding neomers were also found in other known cancer-associated genes, such as tumor protein p53 (*TP53*), B-Raf proto-oncogene serine/threonine kinase (*BRAF*) and phosphatidylinositol-4,5-bisphosphate 3-kinase catalytic subunit alpha (*PIK3CA*). The most frequent neomer was located in a noncoding region, within the telomerase reverse transcriptase (*TERT*) promoter, which is known to be associated with numerous cancer types^27^ and poor prognosis^28^. This mutation, called -124C>T or C228T, is extremely common in numerous cancer types^29^ and is thought to disrupt a G-quadruplex^30^ leading to the binding of GAPB^31^, an ETS transcription factor, resulting in increased *TERT* expression. We found this mutation in 97 patients, with the highest incidence in glioblastoma (51%), fitting with its known high prevalence rate and diagnostic use for this cancer type^32^.

**Table 1.**
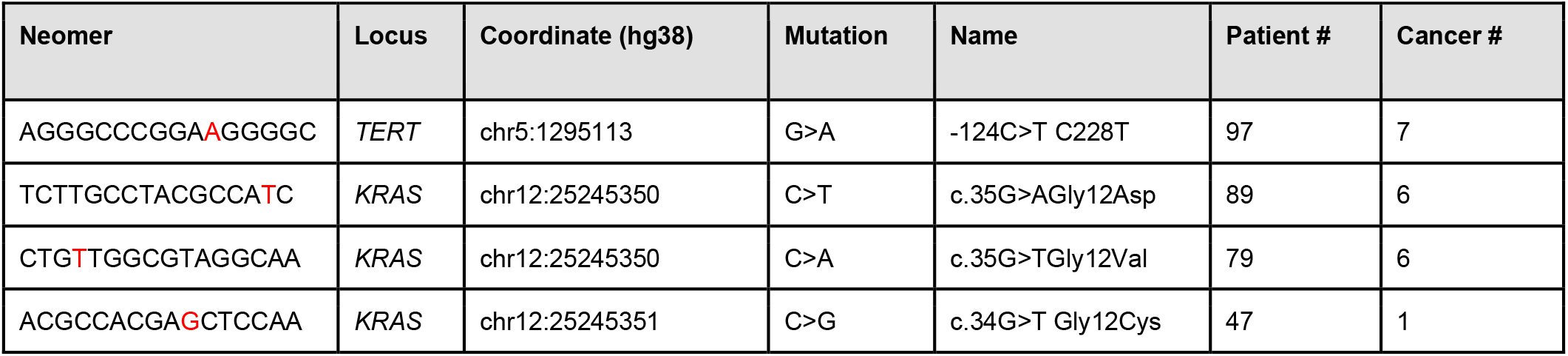
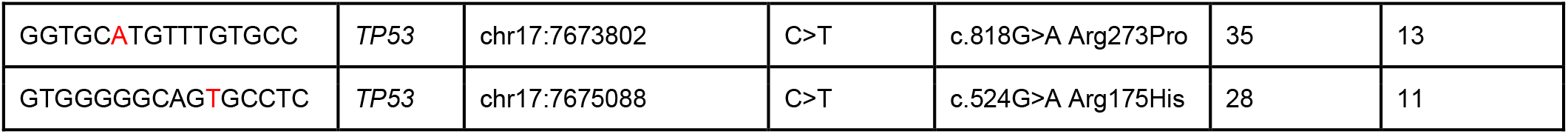
Common cancer-associated neomers. Six of the most common neomers created by a single mutation. The nucleotide in red is the neomer causing mutation.

We also identified several neomers that are frequently created by different mutations (**Table 2**). Interestingly, some of these frequently recurrent neomers are created by different mutations, yet are predominantly found in one cancer. For example, GTTTTTCTCCTAGACC is found 40 times in skin cancer at 31 different loci while CTGGCAGTGAGCCACG is found 21 times in liver cancer across 18 loci. The majority (98%) of these frequent neomers reside in noncoding regions, and many of them reside in intronic regions (35%). For example, CGACGTTCTGCCCACT is found in 32 loci, primarily in pancreatic and stomach cancer. Of those loci, 21/32 (65.6%) were found in noncoding regions nearby pancreatic cancer associated genes. These include, for example, the C-C motif chemokine ligand (*CCL4*)^33^, the POM121 transmembrane nucleoporin like 12 (*POM121L12*), which is commonly mutated in gastrointestinal cancers^34^ and the potassium voltage-gated channel modifier subfamily V member 1 (*KCNV1*) gene, where promoter hypermethylation has been associated with both pancreatic^35^ and esophageal cancer^36^.

**Table 2.**
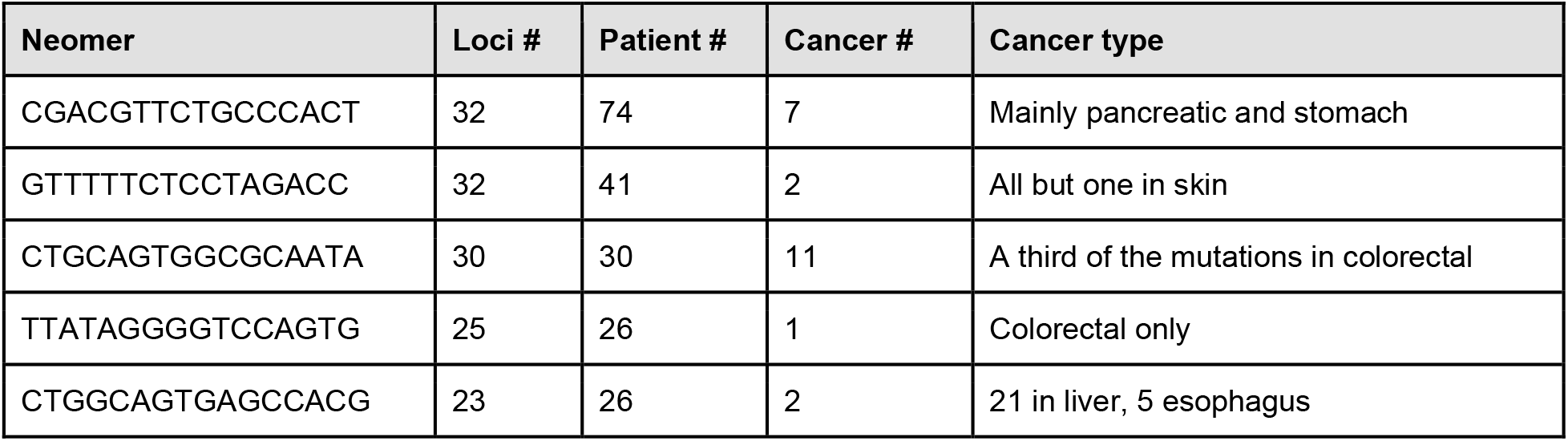
Frequent cancer-associated neomers. Five of the top frequently recurring neomers created by several different mutations.

To further validate that nullomers can detect cancer-associated mutations, we examined whether they are enriched in known cancer driver mutations. We annotated a cohort of driver mutations from 29 different cancer types^37^ for their overlap with 16bp nullomers. In the pan-cancer analysis, we identified 19,594,212 nullomers resulting from 50,167 putative driver mutations (**Fig. 1f, Table 1**). For specific cancer types, we found that on average, 81% of driver mutations resulted in one or more nullomer, ranging between 63.88% and 86.50% in pancreatic and lung cancer respectively (**Extended Data Table 1**). The number of nullomers also varied by tissue type, ranging between 92 in thyroid and 9,434 in skin (**Fig. 1d**). In general, driver mutations were 1.4-fold more likely to result in the creation of a nullomer (permutation test, p-value<0.001). Taken together, our results suggest that a large subset of clinically relevant cancer mutations are associated with neomers, i.e. recurrent nullomers.

### Generation of a cancer subtype neomer classifier

We next set out to assess whether neomers can be used to distinguish between cancer types. We filtered neomers by keeping only those that appeared >=*ri* times in specific cancer type *i* (**Supplementary Table 1**). Comparison of the set of neomers associated with each cancer type revealed a small overlap, as indicated by the Jaccard index which is <0.04, suggesting that each cancer type has a distinct neomer signature (**Fig. 1e**). We also counted the number of times neomers are found in each patient, finding that patients are strongly enriched for only one set of cancer specific neomers (**Fig. 1f**).

We then tested if our annotated cancer-specific neomers can be used to classify tumor samples. We trained a support vector machine classifier to identify tumor type. The classifier takes as input a 21-dimensional vector indicating the number of neomers found for each cancer specific set. Evaluation using 10-fold cross-validation, revealed that our classifier achieves both high sensitivity and specificity, with an F1 score of 0.92 and an accuracy of 0.99 (**Fig. 2a-b**). Performance was better than a recent deep learning mode^l21^ and also requires less computational resources to train as once neomers have been extracted, training and testing takes only a few minutes.

**Fig. 2:**
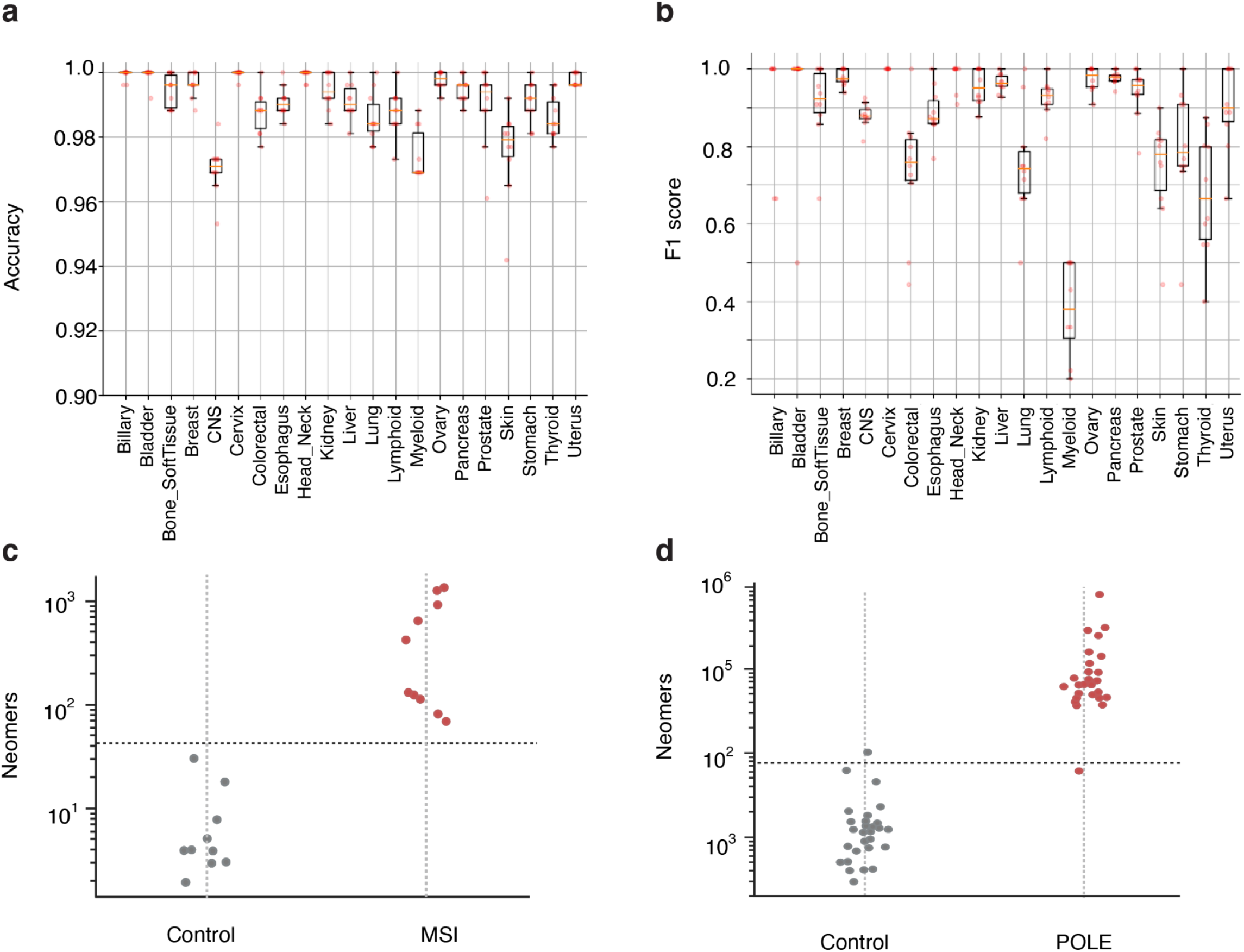
Neomers can distinguish cancer features. **a-b**, Classifier accuracy (**a**) using an unsupervised classifier and F1 (**b**) score for the same classifier for each of the twenty-one cancer types. **c**, Separation of MSI and MSS samples using a supervised selection of neomers. **d**, Separation of POLE proficient and deficient samples using nullomers. In **c-d** the vertical line displays the harmonic mean.

### Neomers can distinguish additional cancer features

We next tested whether a hand-crafted feature selection approach, i.e. using neomers that are thought to be informative based on prior biological knowledge, would improve performance. For this approach, we initially utilized microsatellite unstable (MSI) and microsatellite stable (MSS) cancers. MSI is associated with better cancer prognosis, increased benefits from surgery and higher sensitivity to immunotherapy, but with a lack of efficacy from adjuvant treatment^38^. Since MSI cancers are associated with mutations at polyA and polyT stretches^39^, we hypothesized that neomers containing these motifs would be able to effectively distinguish these two cancer types. We identified ten MSI samples from a cohort of 560 breast cancers^40^ and compared them to ten randomly selected MSS samples from the same cohort. We found that the polyA/T neomers were able to separate the two categories with an accuracy of 100% (**Fig. 2c**).

We next applied a similar strategy to distinguish patients with DNA polymerase epsilon catalytic subunit (POLE) deficiency, as these tumors are known to respond more favorably to immune checkpoint inhibitors^41–43^. We identified 25 patients from the TCGA dataset labeled as POLE deficient, and searched for neomers created through a TCT>TAT or TCG>TTG mutation, which are the most common types of mutations in this context^43^. Comparing against POLE proficient tumors, we found that the number of neomers identified for each group have very little overlap (**Fig. 2d**), and the classifier achieved an accuracy of 96%.

### Neomers detect early-stage lung cancer

We next tested whether neomers could be used to diagnose cancer in cfDNA. We first used lung cancer as our test case, due to having an ample amount of cancer-associated neomers (**Supplementary Table 1**), difficulty to diagnose using current techniques, with early stages being mostly asymptomatic^44,45^, and being the leading cause of cancer-associated mortality worldwide^44,46^. We initially excluded all neomers that could arise due to common germline variants^19^. For each lung cancer associated neomer, we characterized all possible single nucleotide substitutions in the reference genome that could give rise to this neomer. By intersecting this list of neomer creating substitutions with known germline variants identified by the gnomAD project^47^, we calculated the probability that each neomer will be present in an individual. We excluded all neomers that are found in the population with p>0.05, providing 153,026 lung cancer associated neomers for subsequent cfDNA analyses.

We assembled a cohort of cfDNA from the plasma of 381 individuals. These included 201 lung cancer patients from all stages with a strong emphasis on early stages: 77 stage I (38% of cancer cases), 36 stage II (18%), 34 stage III (17%) and 53 stage IV (27%). The majority of our cases were adenocarcinoma (50.2% of cancer samples), followed by squamous cell carcinoma (37.8%) (**Supplementary Table 2**). The median age was 64+/-7.8 years old, with patients as young as 42 years old. We also had 180 controls, considered cancer free (e.g. “healthy”, no cancer of any kind detected >2 years after sampling) with a median age of 58+/-8.1 years and 90% fitting the criteria for the United States Preventive Services Task Force (USPTF) age interval for early screening for lung cancer. Of the healthy individuals, 21% were active smokers (37.2% were noted as unknown), while 64.2% of the cancer patients were noted as active or past smokers (complying with USPTF guidelines for lung cancer screening)(**Supplementary Table 2**). Samples were paired end sequenced at a genomic depth of 5-10X coverage (*C*) and analyzed for neomers. To reduce the impact of sequencing errors, we only considered neomers that were found on both read-pairs, resulting in 18,090-55,409 read-pairs per sample. To further avoid spurious neomers, only neomers that overlapped at least two read-pairs and overlapped the human genome were used for subsequent analyses. Of note, an important computational advantage compared to conventional mutation calling pipelines is that only a single pass is made across the reads to identify those containing neomers, and only those reads are aligned to the reference genome. Using these data, we trained a logistic regression classifier, achieving an AUC score of 0.93 using 10-fold cross validation and a sensitivity of 92.5% (specificity of 85%) (**Fig. 3a-b**). Importantly, our classifier also performed well for early stages, with an AUC of 0.94 for stage I and sensitivity of 96.1% (specificity of 85%) (**Fig. 3c-d**).

**Fig. 3:**
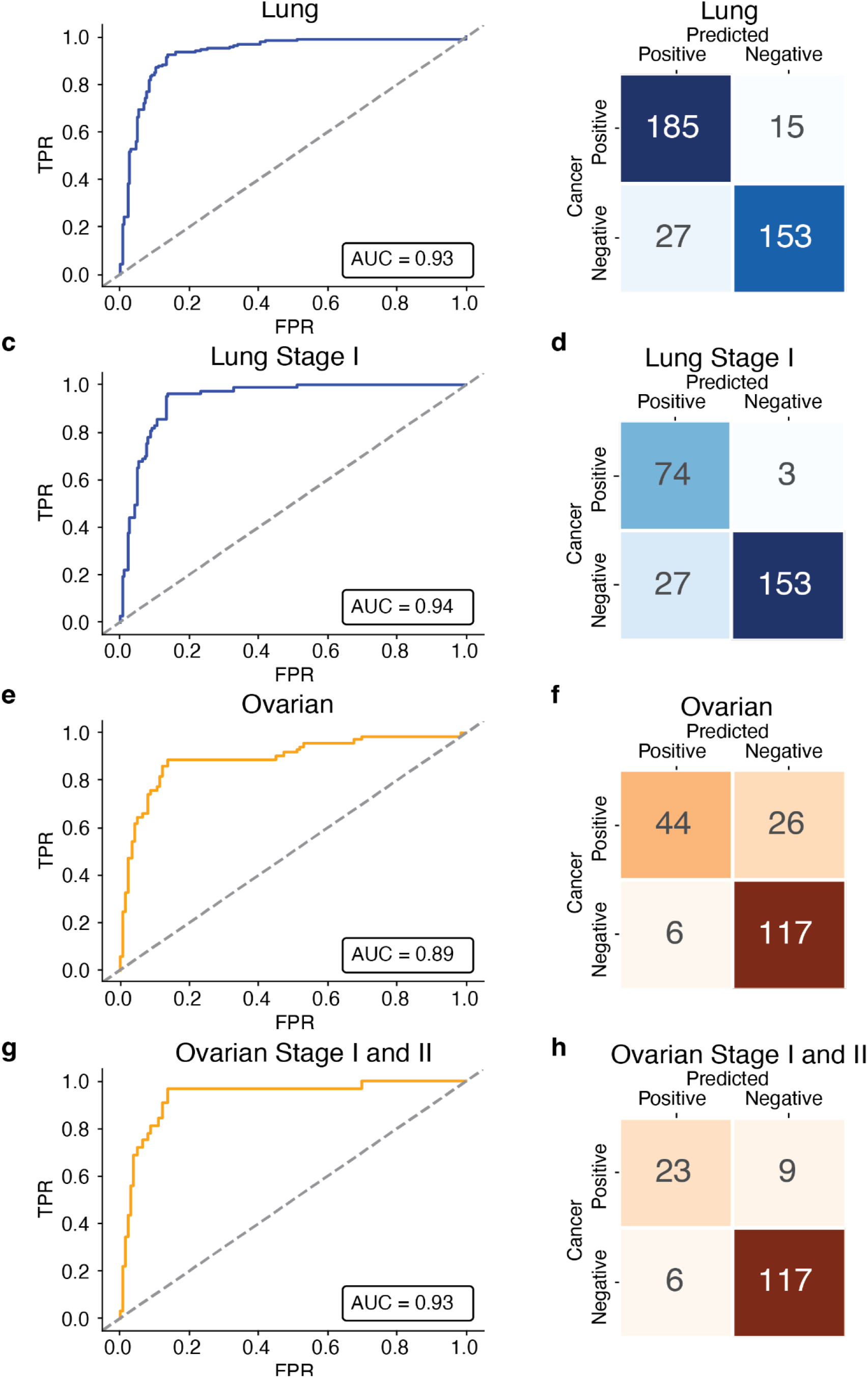
Neomers can detect lung and ovarian cancer from cfDNA. **a-b**, Receiver operator characteristics (ROC) curve (**a**) and confusion matrix (**b**) for the neomer based classifier for cfDNA from 381 lung cancer patients and 180 controls. **c-d**, ROC curve (**c**) and confusion matrix (**d**) from 77 stage I lung cancer cases and 180 controls. **e-f**, ROC curve (**e**) and confusion matrix (**f**) for ovarian cancer from 70 cases and 123 controls. **g-h**, ROC curve (**g**) and confusion matrix (**h**) for 24 stage I and 8 stage II ovarian cancer and 123 controls.

### Neomers detect early-stage ovarian cancer

We next set out to test how our neomer classifier performs with a low amount of tumor-associated neomers. We focused on ovarian cancer which only has 12,788 associated neomers (amongst our analyzed TCGA WGS datasets) (**Supplementary Table 1**). In addition, as ovarian cancer does not have an effective screening test^22^, most women are diagnosed at stages IIIC and IV, when 5-year survival rates are 39% and 17%, respectively^23^, reflecting a pressing need for early detection methods. We obtained plasma samples and generated cfDNA from 70 patients (**Supplementary Table 2**) with a median age of 57+/-16.1 years. Our analyzed cohort is comprised of 45.7% early stage samples (24 stage I and 8 stage II), and the rest from late stage (34 stage III and 4 stage IV). Samples were similarly paired end sequenced at a genomic depth of 5-10X coverage (*C*) and analyzed for neomers. As controls, we used female donors from our control donor group, totaling 123 control cfDNA samples with a median age of 58+/-7.9 years (**Supplementary Table 2**). Using the same classifier as before, we obtained an AUC of 0.89 and a sensitivity of 62.86% with a specificity of 95.12% for all stages (**Fig. 3e-f**). Importantly, for early stages (stage I and II), we obtained an AUC of 0.93 and sensitivity of 71.9% with specificity of 95.12% (**Fig. 3g-h**). Combined, these results suggest that neomers can detect cancer even with a low tumor mutation burden and more importantly could potentially be used to diagnose early-stage ovarian cancer for which there is currently no available detection method.

### Neomers alter promoter activity

Only a small number of mutations in gene regulatory elements have been found to be associated with cancer^24,48,49^. As our results show that driver mutations are enriched for neomers (**Fig. 1d**) and the majority of our neomers (98%) lie in noncoding regions, we hypothesized that neomer creating mutations in noncoding regions could be used to detect cancer-associated mutations in gene regulatory elements that have a functional consequence. Of note, the top patient recurrent neomer was in the *TERT* promoter (**Table 1**), which is associated with numerous cancers^27^. We chose to focus on prostate cancer since it is associated with a relatively small number of neomers (N=4,621; median per patient=29.5), allowing us to characterize all the detected neomers for this cancer via MPRA in the following section. Moreover, 80-90% of prostate cancer is dependent on Androgen Receptor (AR) signaling^50^ allowing us to use LNCaP-FGC cells, which are androgen-dependent, for our reporter assays. We initially set out to test neomers found in promoters for luciferase reporter assays (**Fig. 4a**) using the following criteria: i) neomers that reside in a promoter based on ENCODE annotations^51^; ii) the gene regulated by the promoter is associated with prostate cancer. Our list (**Supplementary Table 3)** included neomers in: 1) a promoter between two divergent genes, *RPS2* and the lncRNA gene *SNHG9* (**Fig. 4b**), both of which are overexpressed in prostate cancer^52^; 2) a promoter between two divergent genes, *TMEM127* and *CIAO1* (**Fig. 4c**), with the former being downregulated in prostate cancer^53^; 3) a promoter between two divergent genes, *TTC23* and *LRRC28*, with the former showing aberrant splicing that relates to therapy resistance in prostate cancer cells^54^; 4) The promoter of *GNAI2*, a protein that interacts with *CXCR5*, which positively correlates with prostate cancer progression^55^; 5) A promoter between two divergent genes, *PRICKLE4* and *FRS3*, with the latter thought to affect malignant but not benign prostate cells^56^.

**Fig. 4:**
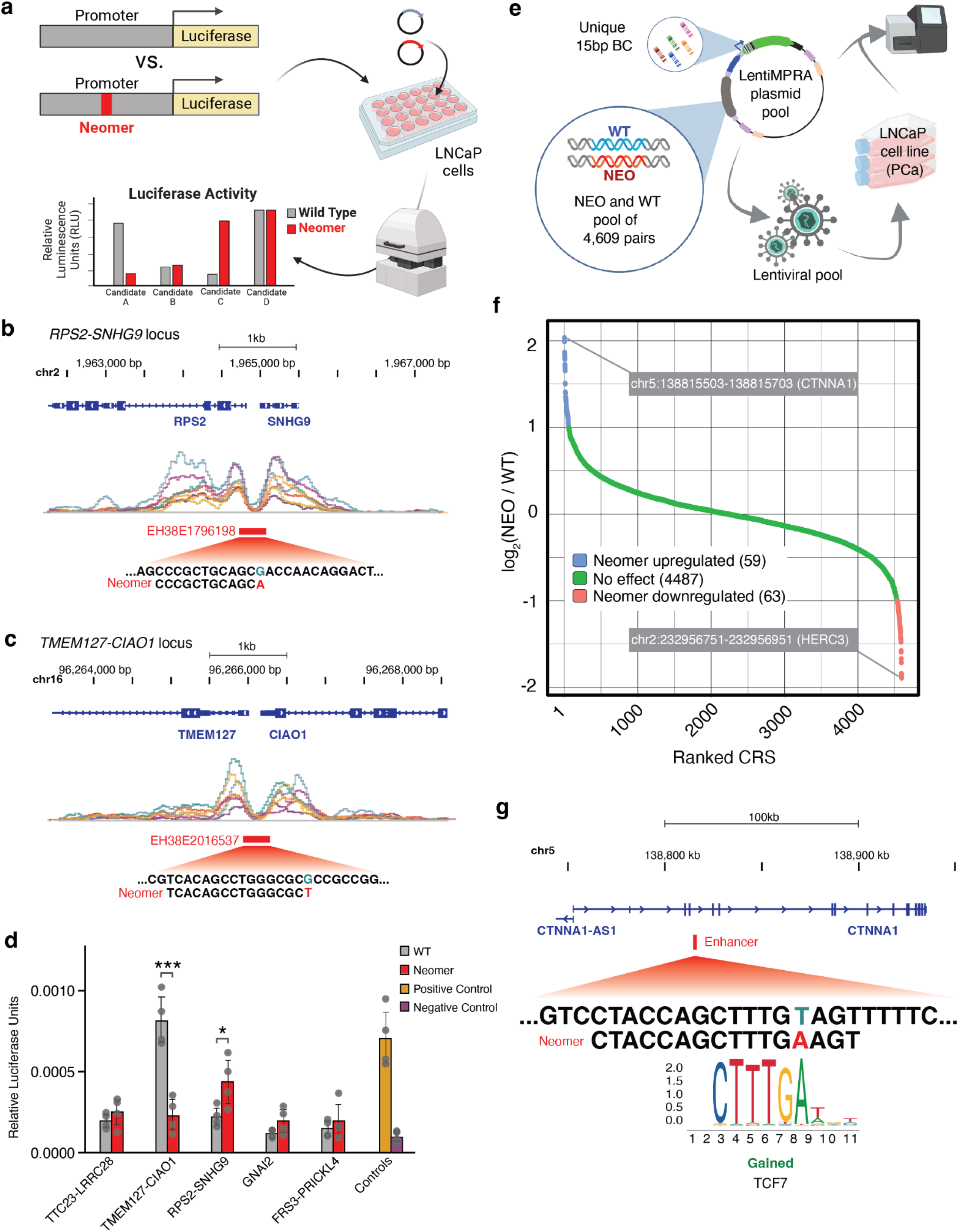
Neomers alter the activity of gene regulatory elements. **a**, Schematic describing the luciferase reporter assay. **b**-**c**, Integrative Genomics Viewer track snapshot showing the location of a prostate cancer associated neomer in the promoters of *RPS2*-*SNHG9* (**b**) and *TMEM127*-*CIAO1* (**c**). The top track shows the gene location (GENCODE V36), beneath it the ENCODE layered H3K27ac marks in this region, below that the promoter location (red line) according to ENCODE cCRE annotations and the most lower track shows the neomer sequence. **d**, Relative luciferase units from a luciferase promoter assay containing either the wild-type (WT) or nullomer variant. Transfection efficiency was normalized using Renilla luciferase and significance is calculated using a two-way ANOVA with multiple testing and Šidák correction. **e**, Schematic showing the lentiMPRA workflow testing 4,906 wild type (WT) and neomer (NEO) sequences. **f**, The regulatory impact of 4,609 neomer creating mutations, presented as log2 fold change of neomers containing sequence over wild type sequnce (x-axis) and ordered from highest to lowest (y-axis). **g**, A neomer in the *CTNNA1* locus had the highest activating score and is postulated to lead to a gain of a TCF7 motif, as predicted by FIMO^57^.

We cloned the promoter sequences with and without the neomer into a luciferase promoter assay vector and compared their activity in LNCaP cells (**Fig. 4a**). Two out of the five assayed promoters showed a significant effect on reporter activity between the neomer and wild type allele (**Fig. 4d**). For the *RPS2*-*SNHG9* promoter, the neomer led to significantly increased activity, in line with this gene being overexpressed in cancer^52^. For the *TMEM127*-*CIAO1* promoter, the neomer completely abolished activity, fitting with its observed downregulation in prostate cancer^53^. Combined, our experimental results show that neomers could have a significant effect on promoter activity and could potentially be used to identify cancer associated gene regulatory mutations.

### Neomers alter enhancer activity

We next set out to test in a high-throughput manner whether neomers could be used to identify mutations that affect the function of active gene regulatory elements, such as promoters and enhancers. We annotated all the prostate-associated neomers that reside in noncoding regions, obtaining 4,609 loci and generated a lentivirus-based MPRA (lentiMPRA) to test the regulatory activity with and without the neomer of all these sequences (**Figure 4e**). Two hundred base pair sequences, where the position of the neomer is used as a center, were synthesized and cloned upstream of a minimal promoter followed by a GFP reporter gene. Lentivirus was generated and the library was infected using three technical replicates into LNCaP cells and RNA and DNA barcodes were sequenced to determine regulatory activity, as previously described^58^. We observed a good correlation between RNA/DNA ratios (Pearson 0.72-0.77; **Extended Data Fig. 3a**) as well as for DNA and RNA barcodes separately (**Extended Data Fig. 3b-c**) across all three technical replicates. Out of the 4,609 loci, 567 showed significant regulatory activity due to the neomer (RNA/DNA ratio >= 1.5). Amongst them, 94 neomers and wild type sequence pairs showed significant differential enhancer activity, with 48 having increased activity due to the neomer (NEO/WT >= 2) and 46 decreased activity (NEO/WT<= 0.5) (**Fig. 4f, Supplementary Table 4**).

The neomer that showed the highest level of increased activity compared to reference (4.09-fold) is located in an intron of the catenin alpha 1 (*CTNNA1*) gene. This gene is a core member of the cadherin/catenin complex and is involved in the regulation of the Wnt/beta-catenin pathway, which has been widely studied in cancer^59^, including prostate cancer^60^, and is the target of several therapies^61,62^. TFBS analysis of this neomer using FIMO^57^ and JASPAR^63^ found that it leads to a gain of a TCF7/TCFL1 motif (MA0769.2) (**Fig. 4g**), which is known to play a role in prostate cancer malignancy^64,65^. A neomer residing in the third intron of the HECT and RLD domain containing E3 ubiquitin protein ligase (*HERC3*) gene led to the lowest downregulation of reporter activity (2.7-fold; **Fig. 4f**). HERC3 is an androgen receptor target and its dysregulation has been associated with epigenetic modulations and poor outcome in prostate cancer^66,67^. In addition, for colorectal cancer, this gene was found to be downregulated, is known to inhibit metastasis and is associated with overall lower survival^68^. Taken together, our MPRA results suggest that neomers could be utilized to identify gene regulatory mutations in cancer.

## Discussion

Cancer is a DNA mutation associated disease. Here, we show that by analyzing cancer WGS, both from tumors and cfDNA, we can find cancer-associated DNA mutations that lead to the generation of neomers, short sequences that are predominantly absent from genomes of healthy individuals. Further analyses of these sets of neomers show that they can be used not only to classify cancer tissue of origin, but also to identify additional cancer features, such as MSI or POLE deficiency with high accuracy. Analysis of cfDNA WGS generated from lung and ovarian cancer patients, finds that neomers could be used to detect cancer, including in early stages, with high specificity and sensitivity. This is particularly important for ovarian cancer, for which there is currently no effective screening test^22^ with most women being diagnosed at later stages when survival rates are low^23^, we were able to detect early stage (stage I and II) cancer with an AUC of 0.92. Finally, using promoter assays and MPRA, we show that neomers can be used to detect cancer-associated mutations that have a functional effect on regulatory element activity.

We utilized 2,577 patients from 21 different cancerous tissues to develop a cancer tissue of origin classifier. Overall, we observed almost no overlap between neomers found in different cancer types. This allowed us to detect cancer tissue of origin with extremely high specificity and accuracy (F1 score of 0.92 and an accuracy of 0.99) performing better than recent deep learning models^21^. In general, the classifier has better performance for cancer types with more patients and high mutation burden (**Fig. 2A-B**). Our analyses also showed that other than tissue origin, nullomers can also be used to detect additional cancer features. Future work that will utilize additional cases and meta data would allow to test whether nullomers and neomers can diagnose additional tumor features and also detect other cancer characteristics such as chance of recurrence, predict and/or monitor response to treatment, disease-free survival and overall survival.

For cfDNA, despite various known challenges, including fragmentation and low levels of ctDNA, we obtained high AUC scores, and our results for lung cancer compare favorably with a recent study based on metabolite profiling^69^. Encouragingly, this holds also for early-stage lung cancer. For ovarian cancer, which is associated with a low tumor mutation burden and has hitherto been extremely challenging to detect at early stages, we managed to detect, even with low sample numbers (N=70), both late and more importantly early stages (stage I and II), obtaining an AUC of 0.93 and sensitivity of 71.9% with specificity of 94.44%. Since the number of neomers increases with the number of tumor samples profiled, obtaining additional WGS datasets from tumors and matched controls would be beneficial in improving the ability of our classifier to diagnose both these and other cancers.

Our cfDNA detection approach has several advantages over current methods: 1) Detecting short DNA sequences that are enriched in cancer samples provides an easy to use diagnostic that could allow detection from low amounts of ctDNA. In addition to sequencing-based assays, alternate techniques could potentially be used, such as CRISPR-based detection tools that utilize Cas12 or Cas13^70^, that can also allow the testing of thousands of sequences in parallel^71^. In addition, with neomer-based diagnostics potentially not needing large amounts of starting material, cfDNA could be collected from urine, sputum, saliva or other bodily fluids, which were shown to be a viable but reduced source of cfDNA^72,73^. 2) Our WGS approach is applicable to more sparsely sequenced samples. We carried out a downsampling analysis and found that already at 3x coverage we obtain similar accuracy for both lung and ovarian cancer (**Fig. S2**). 3) As our diagnostic is based on neomer detection followed by genome alignments of nullomer encompassing reads, it is extremely effective from a computational standpoint. 4) Neomers could easily be combined with other sequence or analyte based cancer biomarkers and risk factors to improve the diagnostic positive predictive value. For example, it was shown that combining a blood test that detects both protein biomarkers and DNA mutations along with positron emission tomography - computed tomography (PET-CT) could detect multiple cancers^74^. In another example, the use of cfDNA fragmentation patterns combined with CT imaging, clinical risk factor and serum levels of carcinoembryonic antigen significantly increased the ability to diagnose lung cancer^75^. Adding neomers to known cancer-associated coding mutations in the screening of cfDNA could also increase sensitivity and specificity. In summary, coupling neomer-based diagnostics to existing cancer biomarkers and risk factors could improve the power to detect various cancer subtypes.

As nullomers/neomers do not exist in the human genome they could also be exceptional candidates for neoantigens, to be targeted via immunotherapy. Previous work has shown that minimal absent words, short sequences that are absent from a genome or proteome, could be used to identify phosphorylation sites of high confidence, some of which could be associated with cancer^76^. Analysis of the Immune Epitope Database of validated antigens^77^ found that 13 of the recurrent coding neomers can create neoantigens with predicted strong binding levels that were subsequently validated (**Supplementary Table 5**). From the 1,700 neoantigens with strong binding levels, only 1.72 (p-value<1e-8, hypergeometric test) is expected to correspond to a neomer, suggesting that missense mutations also resulting in neomers are 7-fold more likely to also generate strongly binding neoantigens.

Neomers can be used as a novel tool to identify cancer-associated gene regulatory mutations. Amongst the 210 prostate cancer promoter neomers, we selected five promoters and found that two of them significantly affected promoter activity due to the neomer. Their difference in activity was in line with the gene’s expression change in prostate cancer, with *RPS2*-*SNHG9* having increased activity fitting with *RPS2* overexpression in prostate cancer^52^ and *TMEM127*-*CIAO1* abolishing activity, in line with *TMEM127* observed downregulation in cancer^53,78^. Our MPRA library of 4,609 neomer causing mutations in enhancers revealed that 2.6% can change gene expression by >1.5-fold, suggesting that a subset of these mutations could have important functional consequences in cancer. Understanding tumor onset and its transition to a metastatic cancer has been limited mostly to coding genes or their pre-determined regulatory sequences^79–81^. With the vast majority of mutations falling into non-coding regions, making them much more challenging to interpret, novel approaches are required for identifying mutations that have functional consequences. Here, we show that by prioritizing mutations that result in neomers, it is possible to shortlist candidates that are likely to have a phenotypic impact. This approach can be applied to any cancer type and thus holds the potential to expand our catalog of functionally relevant cancer driver non-coding mutations.

In summary, we show that neomers can provide a powerful tool for cancer diagnosis. As they can easily be detected via sequence or CRISPR-based tools, it should be straightforward to integrate them in current routine cancer diagnostic tests and their use could increase the sensitivity and specificity of these tests. Combining neomer-based screening with clinical characteristics and additional diagnostic tools and features could increase the positive predictive value. In addition, as cfDNA could also be isolated from urine and saliva, and detection of these sequences only requires a relatively small amount of DNA, neomer-based diagnosis could be carried out in a non-invasive manner. Our work also suggests that neomers could be used to highlight cancer-associated gene regulatory mutations which have been difficult to identify. Further high-throughput characterization of these mutations could allow the detection of bona fide cancer-associated functional regulatory mutations that could be used for diagnosis and treatment.

## Methods

### Computational characterization of nullomers

The GRCh38 reference assembly of the human genome was used throughout the study. Nullomer extraction was performed for kmer lengths up to 17 base pairs using the algorithm described in our previous work^19^. The reverse complement of a nullomer will also be a nullomer and throughout this manuscript when counting nullomers, the reverse complement of nullomer *i* was also considered separately, unless *i* is a palindrome. Substitutions and indels identified from WGS of tumor samples from 2,577 individuals across 21 tissues were obtained from https://dcc.icgc.org/releases/PCAWG/^24^. Recurrent nullomers (neomers) (*ri*) were annotated as those that resulted from substitutions or indels across two or more patients within a cancer type. When possible, *ri* was chosen to get ∼10,000 neomers from each tissue, otherwise it was set to 2 (**Supplementary Table 1**). Driver mutation-derived neomers were defined as nullomers detected from driver mutations and were identified using the catalogue of driver mutations derived from the intOGen database^37^, for which we grouped driver mutations by tissue of origin.

### Classification of tumor tissue of origin using neomers

We trained a classifier to distinguish classified tissue of origin for a cancer sample based on observed neomers using the libSVM package for Julia^82^ with default parameters to train a support vector machine classifier with a linear kernel. We used 10-fold cross validation whereby the classifier was evaluated on a held-out fraction of the data. The set of neomers for each cancer type was recalculated for each round to only include the patients in the training set.

### Supervised selection of nullomers

The MMR status of each biopsy sample was derived from^83^. The model was trained on neomers identified in MSI samples and the performance of the algorithm evaluated. For the MSI versus the MSS samples, we counted the number of neomers that contained either AAAAAAAA or TTTTTTTT repeats, since MSI cancers have been associated with mutations of polyA/T repeats ^39^. The threshold for determining MSI or MSS was set as the harmonic mean of the maximum number of counts in the MSS set and the minimum number of counts in the MSI set. The POLE deficiency status of each biopsy sample was derived from^83^ and we used a similar strategy to that of MMR status, but instead counted neomers created through either a TCT>TAT or TCG>TTG mutation. Since the number of patients in each category was limited, we used a 5-fold cross validation. For ovarian cancer analyses, two sets of nullomers were used. From the nullomers of length *k=13* identified in^19^, we retained only the ones that could not be created by any of the single base-pair substitutions identified in gnomAD v2^47^. Similarly, we selected all nullomers of length *k=15* of order 1, i.e. nullomers that can only be created from the human reference genome through at least two insertions, deletions, or substitutions.

### cfDNA extraction and WGS

For lung and ovarian cancer, samples were purchased from ProteoGenex Inc. (Inglewood, CA, USA) and IndivuMed GmbH (Hamburg, Germany) and also obtained from the labs of K. Syrigos (Sotiria Hospital, National and Kapodistrian University of Athens, School of Medicine, Athens, Greece), J. Chapman (UCSF, San Francisco, USA), J. Witte (UCSF, San Francisco, USA) and C. Maher (WashU, St. Louis, USA). All human sample work was carried out under the approved UCSF human research protection program institutional review board protocol number 21-35920. cfDNA was extracted from 1mL of plasma, following centrifugation at 4C at 600 rpm for 3 minutes, to remove larger debris, using the QIAamp Circulating Nucleic Acid Kit (Qiagen). cfDNA was eluted in 50uL of elution buffer and measured using the Qubit High-Sensitivity dsDNA kit, and validated for size distribution (160-180bp) using an Agilent BioAnalyzer 2100 Sensitivity DNA chip. Up to 10ng of cfDNA was used for sequencing library construction using the library preparation enzymatic fragmentation kit 2.0 (Twist Bioscience) adjusted with IDT’s xGen UDI-UMI 96 barcodes system (IDT) to replace the Twist universal adapter, and using KAPA HiFi polymerase instead of the polymerase provided by the kit. Libraries for lung, ovarian and their respective controls were sequenced as PE150 using a NovaSeq 6000 S4 system (Illumina) leveraging UMIs to remove PCR duplicates, aiming for 5-10X coverage per multiplexed library, via Novogene.

### Neomer identification in cfDNA samples

To improve the detection of lung cancer mutations in cfDNA, we used neomers that are present in >=2 patients rather than >=3 as was used for the tissue-of-origin classifier. To filter out common population variants, we obtained variant information from the gnomAD v2^47^ and excluded all neomers that were generated due to variants with a frequency >0.05. Variants that were not single base-pair substitutions were not considered. The FASTQ files were scanned for neomers by searching for exact matches to the 16-mers of interest. To reduce errors, only reads where the nullomer was found in both pairs were kept. The remaining reads were then mapped to the reference genome (hg38) using Bowtie2 ^84^ with parameters “--mp 3”. The aligned reads were then analyzed using Samtools mpileup (http://www.htslib.org/) and only those loci that contained >*C*/1.5 read pairs, where *C* is the coverage for the sample, were counted as bona fide neomers. For the analysis of the more deeply sequenced prostate samples (**Fig. 3g**), we adjusted the threshold to *C/10*. When processing the data, not all reads were mapped to the genome, which means that the coverage estimates are based on the total length of the sequenced reads rather than on the mappable reads and should thus be considered an upper bound. To evaluate the ability to detect cancer from cfDNA, we first randomly split samples into training and validation sets. Prior to the split, samples were randomly removed from either the controls or patients to ensure that the two groups were of equal size. From the training set we determined a threshold, *t* = max (number of neomers in control samples from the training set) - 1. Samples in the validation set were then designated as having cancer if the number of neomers detected was >=*t*. To calculate F1 scores, this procedure was repeated 100 times. For the downsampling analysis, each read pair in sample *i* was retained with a probability of *c/Ci*, where *c* is the target coverage and *Ci* is the full coverage of the sample, i.e. sampling without replacement. For each value of *c* the downsampling was repeated 100 times.

### Classification model to detect cancer patients from cfDNA

Neomers were identified for each donor and combined as a neomer density x sample matrix, dij = nij/Cj, where nij is the count of neomer i in sample j and Cj is the sample coverage. Neomer-based classification models were generated to examine the ability of neomers to detect cancer for lung, colorectal, ovarian, pancreatic and prostate cancers. We tested PCA and feature hashing to deal with the sparse nature of our dataset and used feature hashing with 8,192 features as the best performing method. We evaluated five classification models; random forest, linear and radial basis function support vector machines and logistic regression models with L1 and L2 regularization, each evaluated with a 10x10-nested cross-validation. Our benchmarks indicated logistic regression with an L2 penalty as the best performing model across all cancer types. The results obtained were also examined regarding tumor staging for colorectal and lung cancers, in which we had sufficient sample sizes.

### Promoter luciferase assays

Promoter sequences with and without the neomer (**Supplementary Table 3**) were synthetically generated and cloned into the modified Promega promoter assay luciferase vector pGL4.11b (a gift from Dr. Rick Myers, HudsonAlpha) by BioMatik Inc and Sanger sequence verified. LNCaP cells were plated at an initial density of 2*10^5 cells/well in 24-well tissue culture plates and maintained in RPMI medium, 10% FBS supplemented with L-Glutamine and Penicillin/Streptomycin. Plasmids together with a renilla expressing plasmid, pGL4.74 (Promega), at a ratio of 10:1 luciferase:renilla were transfected using the X-tremeGENE™ HP DNA Transfection Reagent (Roche) using 1:4 ratio of DNA (ug) to reagent (ul). 72 hours post transfection luciferase and renilla levels were measured using the Dual-Luciferase Reporter Assay System (Promega) following the manufacturer’s protocol using a GloMax Explorer Multimode Microplate Reader (Promega). Luciferase activity was normalized to renilla levels and presented as Relative Luciferase Units (RLU). Statistical analysis was performed using Prism version 9.0.2 (GraphPad). All values were reported as means (AVG) and standard errors (SE). p values < 0.05 were considered statistically significant.

### Massively parallel reporter assays

The lentivirus-based MPRA was performed as described previously^58^. An oligonucleotide library of 230bp long fragments bearing: 1)4,609 sequences with and without a recurring neomer that was found in prostate cancer patients; 2) 100 scrambled control sequences randomly selected from the library. A pool of 230-nt oligos containing each of these 200-nt sequences flanked by 15 base pair primer sequences on either side was synthesized (Agilent Technologies), amplified, and cloned into the lentiMPRA plasmid. Briefly, amplified inserts were cloned upstream of a minimally active promoter (mP), 5’UTR barcode (BC) and EGFP reporter transcript. We aimed for ∼100 BCs per sequence when constructing and extracting the plasmid library. Each insert was associated with a set of BCs using paired-end (PE150) sequencing on an Illumina NextSeq 500(see Gordon et al. Step 83^58^). Lentivirus was produced and titered and used to infect LNCaP cells at an MOI of 50 virus particles per cell. Three days post infection, to remove non-integrating virus, DNA and RNA were extracted from the three replicates and used for library construction and multiplexing for sequencing. All MPRA-related sequencing was performed using an Illumina NextSeq 500 with either PE150 for the CRS-BC association library or PE15 for the DNA/RNA BC count portion of the protocol. MPRA analysis was carried out using MPRAflow^58^.

### Software availability

We generated an easy to use software package that enables performing nullomer cancer analyses from sequence-based datasets. The package is composed of six functions: 1) EnumerateNullomers, which extracts all nullomers of specified kmer lengths in a FASTA sample; 2) ExtractMutationNullomers, which finds all mutations that cause the resurfacing of a list of nullomers; 3) IdentifyRecurrentNullomers, which identifies nullomers that recur in a dataset through mutagenesis; 4) FindAlmostNullomers, which identifies the positions that can create a list of nullomers genome-wide for every possible substitution and single base-pair insertion and deletion; 5) FindNullomerVariants, which removes nullomers that are likely to result from common variants in a user specified variant VCF file; 6) FindDNANullomersFromReads, which performs the identification of nullomers in raw read samples. The package can be found at: https://github.com/Ahituv-lab/Nullomerator and a Read the Docs (https://readthedocs.org/) tutorial provides in-depth details on how to use the software.

### Comparison to validated neoantigens

We downloaded a list of 1,967 validated neoantigens from http://biopharm.zju.edu.cn/download.neoantigen/iedb_validated.zip. Requiring both predicted strong binding and a positive validation, provided 1,700 neoantigens. To evaluate the enrichment of neoantigens corresponding to neomers, we assumed a hypergeometric distribution with 1,700 draws from an urn with 188,659 white balls (total number of neomers) and 186,067,892 black balls (number of nullomers found with lower recurrency than what was required for neomers).

## Data Availability

For the TCGA data, mutation calls are publicly available in: https://portal.gdc.cancer.gov/
Permission to use the data from https://ega-archive.org/studies/EGAS00001003206 was obtained from the DAC after contacting Dr Ellen Heitzer.
WGS sequencing data was submitted to dbGAP. MPRA sequencing data was deposited in the NCBI short read archive (SRA) as Bioproject PRJNA917083.

https://ega-archive.org/studies/EGAS00001003206

## Acknowledgments

We want to first and foremost thank the individuals who participated in this study. This work was supported in part by the Benioff Initiative for Prostate Cancer Research, the UCSF Catalyst award, the UCSF Innovations Ventures Philanthropy Fund and National Human Genome Research Institute grant number UM1HG011966 (N.A). MH was supported by core funding from the Wellcome Trust and core funding from the Evergrande Center.

## Author contributions

I.G.S., O.Y.B., I.M., M.H. and N.A. conceived the study. I.G.S., I.M., C.C., R.E., G.E.P., M.K., M.H. wrote the code, I.G.S., O.Y.B., I.M., C.C, M.M., M.K., M.H. and N.A. performed the analyses and generated the visualizations, O.Y.B, C.C., R.B., J.S., D.L.C., R.E. and J.K. carried out experimental assays, E.M., G.P., J.S.W., C.A.M., F.F., I.V., N.S., E.P., A.C., K.S. and J.C. provided samples, M.H. and N.A. provided resources and supervised the research, I.G.S., O.Y.B., I.M., M.H. and N.A. wrote the manuscript with input from all authors.

## Data availability

WGS sequencing data was submitted to dbGAP. MPRA sequencing data was deposited in the NCBI short read archive (SRA) as Bioproject PRJNA917083.

Reviewer link: https://dataview.ncbi.nlm.nih.gov/object/PRJNA917083?reviewer=ca59f64g0gm2gjotmusqhhpp5d

