## Supplemental Figures for "Leveraging sequences missing from the human genome to diagnose cancer"

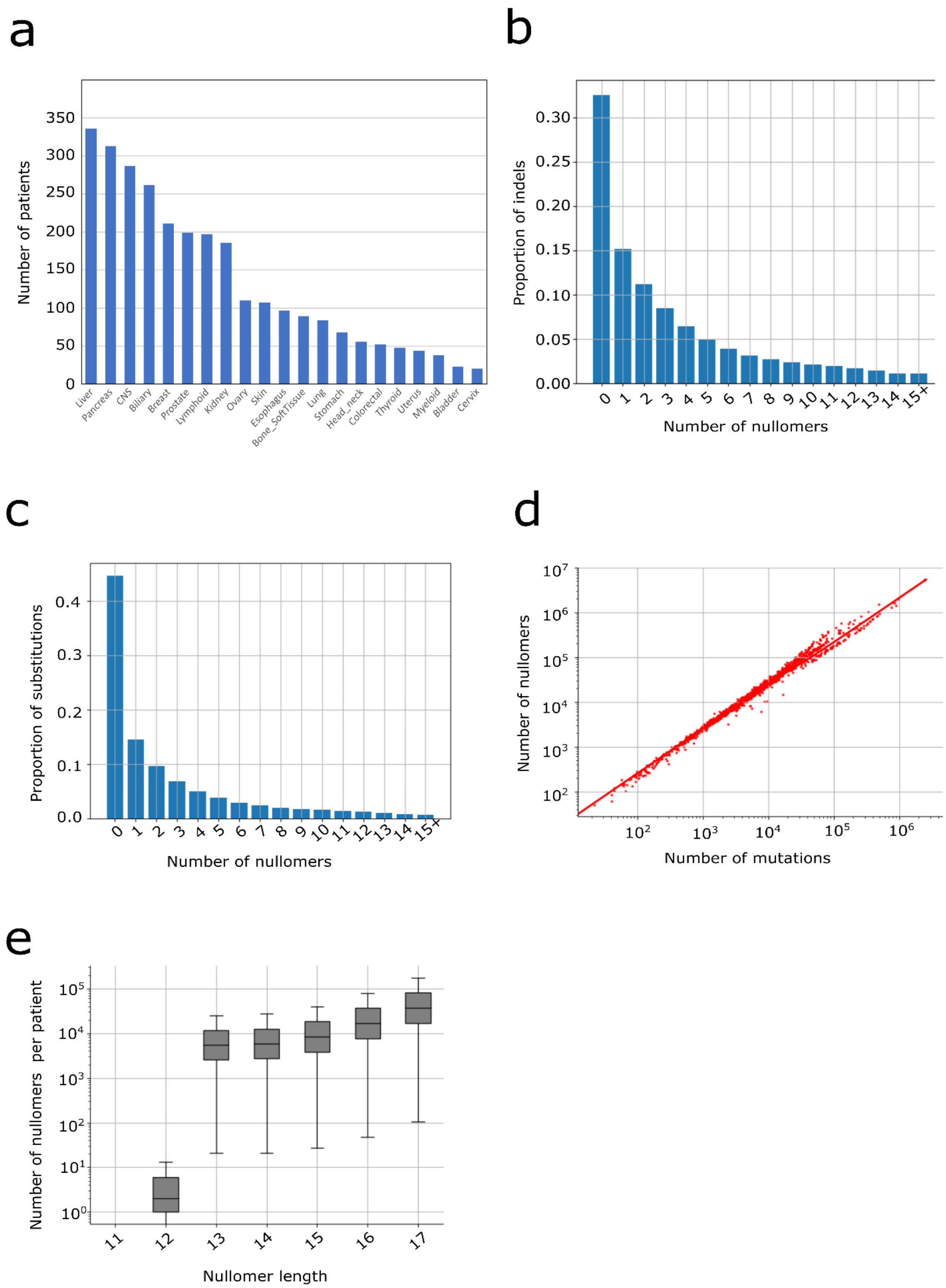


**Extended Data Fig. 1. a,** Number of patients per cancer tissue. **b-c,** Number of nullomers resurfaced due to indels or substitutions observed for each tumor sample (per patient) . **d,** Association between number of mutations and number of nullomers observed. **e,** Number of nullomers of different lengths observed per patient.

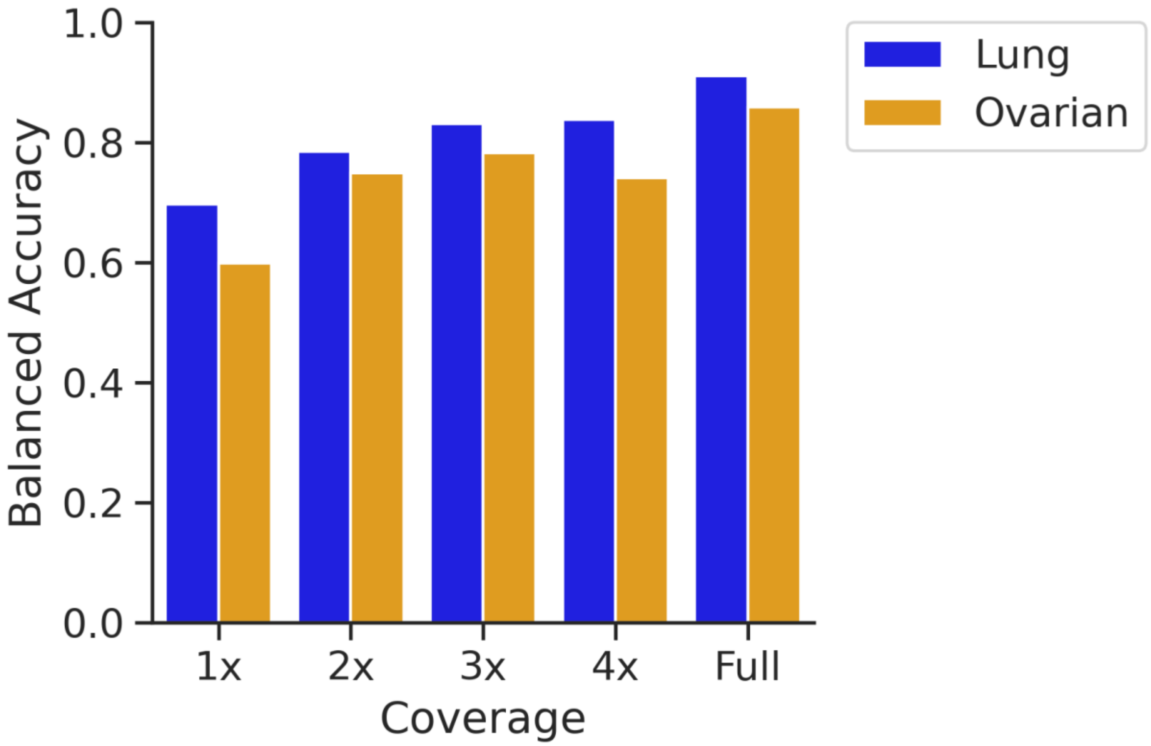


**Extended Data Fig. 2.** Balanced accuracy of lung or ovarian cancer neomer-based assay using different levels of sequencing coverage.

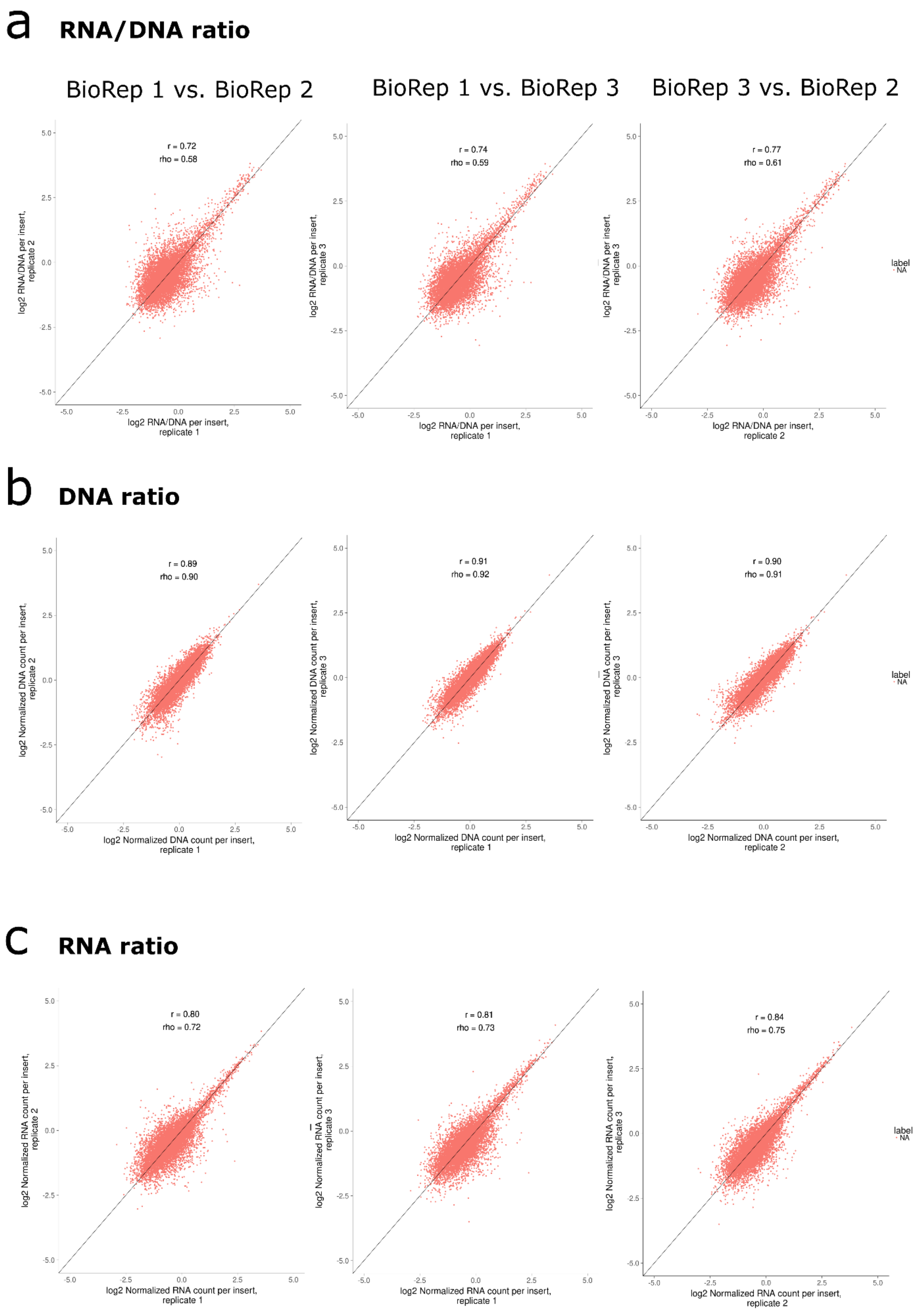


**Extended Data Fig. 3. Correlation between replicates in Porstate MPRA -** All libraries sequences number of normalized reads across all barcodes correaltion between the three biological replicates, as calculated for the RNA/DNA ration (a), DNA barcodes alone (b) and RNA barcode alone (c ).
